# Coarctation duration and severity predict risk of hypertension precursors in a preclinical model and hypertensive status among patients

**DOI:** 10.1101/2023.10.30.23297766

**Authors:** Arash Ghorbannia, Hilda Jurkiewicz, Lith Nasif, Abdillahi Ahmed, Jennifer Co-Vu, Mehdi Maadooliat, Ronald K. Woods, John F. LaDisa

## Abstract

**Background:** Coarctation of the aorta (CoA) often leads to hypertension (HTN) post-treatment. Evidence is lacking for the current >20 mmHg peak-to-peak blood pressure gradient (BPGpp) guideline, which can cause aortic thickening, stiffening and dysfunction. This study sought to find the BPGpp severity and duration that avoid persistent dysfunction in a preclinical model, and test if predictors translate to HTN status in CoA patients.

**Methods:** Rabbits (N=75; 5-12/group) were exposed to mild, intermediate or severe CoA (≤12, 13-19, ≥20 mmHg BPGpp) for ∼1, 3 or 22 weeks using dissolvable and permanent sutures with thickening, stiffening, contraction and endothelial function evaluated via multivariate regression. Relevance to CoA patients (N=239; age=0.01-46 years; median 3.7 months) was tested by retrospective review of predictors (pre-operative BPGpp, surgical age, etc.) vs follow-up HTN status.

**Results:** CoA duration and severity were predictive of aortic remodeling and active dysfunction in rabbits, and HTN in CoA patients. Interaction between patient age and BPGpp at surgery contributed significantly to HTN, similar to rabbits, suggesting preclinical findings translate to patients. Machine learning decision tree analysis uncovered that pre-operative BPGpp and surgical age predict risk of HTN along with residual post-operative BPGpp.

**Conclusions:** These findings suggest the current BPGpp threshold determined decades ago is likely too high to prevent adverse coarctation-induced aortic remodeling. The results and decision tree analysis provide a foundation for revising CoA treatment guidelines considering the interaction between CoA severity and duration to limit the risk of HTN.

## 4 Introduction

Coarctation of the aorta (CoA), a constriction of the proximal descending thoracic aorta, is a common form of congenital heart disease affecting 5,000-8,000 births annually in the U.S^1^. For unknown reasons, treated CoA patients often have a reduced life expectancy from cardiovascular morbidity, primarily hypertension (HTN)^2^.

The blood pressure gradient (BPG) induced by CoA is a primary indication for treatment and is often greatly reduced after surgical resection. Unfortunately, there is a lack of data for the current >20 mmHg peak-to-peak BPG (BPGpp) value in treatment guidelines^3^. The guidelines also reference anatomic evidence of CoA, and a BPGpp >10 mmHg with decreased left ventricular systolic function or significant collateral flow. A 2015 review^4^ called prior evidence (Level C) for this BPGpp guidance suboptimal^5^. Recent guidelines indicate level B-NR^3^, which suggests staying abreast of new information^6^. The >20 mmHg BPGpp seems to date back to surgical outcomes from several decades ago^7^, but persists despite notable advances. Experimental results implementing BPGpp below the current guideline in rabbits has revealed HTN precursors within the proximal region exposed to high blood pressure (BP)^8, 9^. These include medial thickening and stiffening, and endothelial dysfunction, which all persisted after treatment. These data suggest a BPGpp >20 mmHg is not the ideal treatment threshold.

The objectives of this study are to identify the severity and duration of BPGpp that avoids persistent arterial remodeling and endothelial dysfunction using a preclinical model of CoA, and to test whether BPGpp predictors translate to HTN status in treated CoA patients. We hypothesize aspects of CoA including severity and duration can predict whether a patient will be hypertensive since BP is similar and growth curves can be mapped between rabbits and patients^10^.

## 5 Methods

Data supporting the findings of this study are available from the corresponding author upon reasonable request. See the Supplemental Text for detailed methods and a Major Resources Table.

### 5.1 Preclinical model of CoA

Experimental procedures were approved by applicable Animal Care and Use Committees and conformed to the National Institutes of Health Guide for the Care and Use of Laboratory Animals. Briefly, New Zealand white rabbits (total 75; N=5-12/group) ∼10 weeks old and weighing ∼1.0 kg were anesthetized^11^ and randomly designated to surgically undergo discrete CoA of varying severity and associated mechanical stimuli within the proximal descending thoracic aorta by tying suture around the aorta distal to the left subclavian branch against a wire of known diameter (1.6, 2.0 or 2.7 mm). CoA severity depended on diameter of the wire used, resulting in BPGpp within the clinical range (mild: ≤12, intermediate: 13-19, and severe: ≥20 mmHg). The effect of CoA duration was studied using sutures with different dissolving properties (i.e., rapid dissolvable Vicryl, dissolvable Vicryl, or permanent silk) to initiate the stenosis (short: ∼1, long: 3, and prolonged: 22 weeks) at each severity. This resulted in 9 groups (3 severities and 3 durations) as well as a group of non-experimental control littermates. Weekly Doppler ultrasound imaging was performed^12^ until BPGpp estimates via simplified Bernoulli Equation (SBE) no longer changed with body weight. Severity for rabbits in dissolvable CoA groups was estimated noninvasively using Doppler BPGpp evolution curves compared to those from the permanent CoA groups (Figure S1). Rabbits were re-anaesthetized after ∼22 weeks for catheter measurement of simultaneous BP waveforms proximal and distal to the CoA^13^.

Indices of aortic remodeling including thickness, stiffness, and impaired vasoactive response by myography were used as surrogates for HTN relative to the severity and duration of the CoA. Doppler b-mode images were used to quantify wall thickness in the proximal descending thoracic aorta from coarctation-induced mechanical stimuli^8^. Body weight was also measured weekly and thickness evolution was normalized to body weight. Aortas also underwent material characterization by uniaxial extension testing at 37^O^C in an environmental chamber. Resulting stress-stretch curves were used to characterize stiffness. Area under the stress-stretch curve (i.e., strain energy in the sample) was quantified in the stretch range of 1 to 2 as an aggregate measure of overall stiffness. Functional changes were quantified by wire myography of aortic segments (3-4 mm rings). Active function testing was conducted to observe smooth muscle (SM) contraction via phenylephrine (PE) as well as endothelial-dependent relaxation by acetylcholine (ACh) in a half-log increasing dose response from 10^-9^ to 10^-5^ M concentrations. Area under the dose response curves (AUD) was then quantified as an aggregate measure of dysfunction. Detailed results of each index are noted elsewhere^8, 9^ with aggregate alterations and multiple linear regression fits presented below.

### 5.2 HTN in pediatric patients treated for CoA

Electronic medical records of treated CoA patients (N=239) were retrospectively studied after exempt determination by applicable Institutional Review Boards. BPGpp, patient age, sex, height, weight, medication history, and follow-up BP were used to determine BP percentiles and HTN status. Patients <18 years at follow-up were assessed using the Clinical Practice Guideline for Screening and Management of High Blood Pressure in Children and Adolescents^14-16^ implemented via an online calculator^17^. Patients ≥18 years were assessed using Joint National Committee 7 guidelines for systolic/diastolic BP: normal (<120/80 mmHg), pre-HTN (120-139/80-89 mmHg), HTN (includes Stage 1: 140-159/90-99 mmHg and Stage 2: >160/>100 mmHg). Patients with BP percentiles registering as Stage 1 or Stage 2 for systolic or diastolic BP were interpreted as HTN. BPGpp in patient records was determined via SBE. BPGpp noted as “not significant” or “none” were interpreted as zero and average values were used when a range was noted.

### 5.3 Statistical Analysis

Multivariate regression analysis was performed on preclinical rabbit datasets with severity, duration, body weight, and sex as predictors. Predictors were considered relative to aspects of CoA that could impact aortic remodeling in a preclinical model, as well as corresponding indices from retrospective data and review of HTN prevalence in CoA patients^8, 18^. Forward and backward stepwise selection methods were used to ensure the best model was not over-/under-fitting. Three different information criteria, i.e., Akaike information criteria, corrected Akaike information criteria, and Bayesian information criteria, were used to identify the best model^19^. Normality of the residuals were tested through D’Agostino-Pearson, Anderson-Darling, Shapiro-Wilk, and Kolmogorov-Smirnov tests.

To assess the possibility of translating any new BPGpp threshold ranges to patients, predictors from hypertensive CoA patients at follow-up were analyzed from 260 million possible models. Predictors included pre and post-operative estimates of BPGpp (i.e., severity), birth vs surgery date (i.e., duration), follow-up time since surgery, sex, body surface area (BSA) and anti-hypertensive medication (where available). Numerical predictors were scaled by dividing by the corresponding standard deviation so as to make these predictors scale-free and improve comparability of variables^20^. The best model was found with *Glmulti*^21^.

The Classification and Regression Tree was additionally used to model the impact of the predictors on HTN precursors in the preclinical model and HTN status from patient data sets using *Sklearn*^22^. Model predictions were used to compute predictive performance metrics.

Reliability of echo-based measurements was investigated through intraclass correlation coefficient (ICC) by two observers using a random subgroup of rabbit and patient data (n=10).

## 6 Results

### 6.1 Preclinical model of CoA

Table 1 shows sample size, sex, severity and duration of CoA, BP and body weight for rabbits in the study. Aortic thickness and stiffness increased with severity and duration of CoA, whereas active response was impaired. Significant correlation to CoA severity and duration was observed via multiple linear regression fit. Figure 1 shows model prediction vs scatter plot of functional assessment indices at various severities and durations of CoA.

**Figure 1.** Preclinical surrogates of hypertension as a function of CoA severity and duration. All metrics were quantified at the thoracic aorta proximal to the CoA where elevated BP was experienced. Multi-variate regression modeling was performed to predict (A) normalized thickness [μm/kg], (B) stiffness [MPa], (C) contraction impairment [nondimensional], and (D) relaxation impairment [nondimensional] as a function of severity, duration, body weight, and sex. Severity was quantified through trans-coarctation peak-to-peak blood pressure gradient (BPGpp) and duration was quantified in weeks. Logarithmic transformations were used to improved model fits. Specifically, log_2_(BPGpp+12) and log_2_(Duration+12) were used. Reported severities and durations on the axes were transformed back to measured dimensions. Short, long, and prolong classifications are based on sutures used to create CoA for ∼1, 3, and 22 weeks, respectively. Glmulti identified severity, duration, or their interaction as the main effect for each metric of aortic dysfunction. Residuals followed a normal distribution.

**Table 1.**
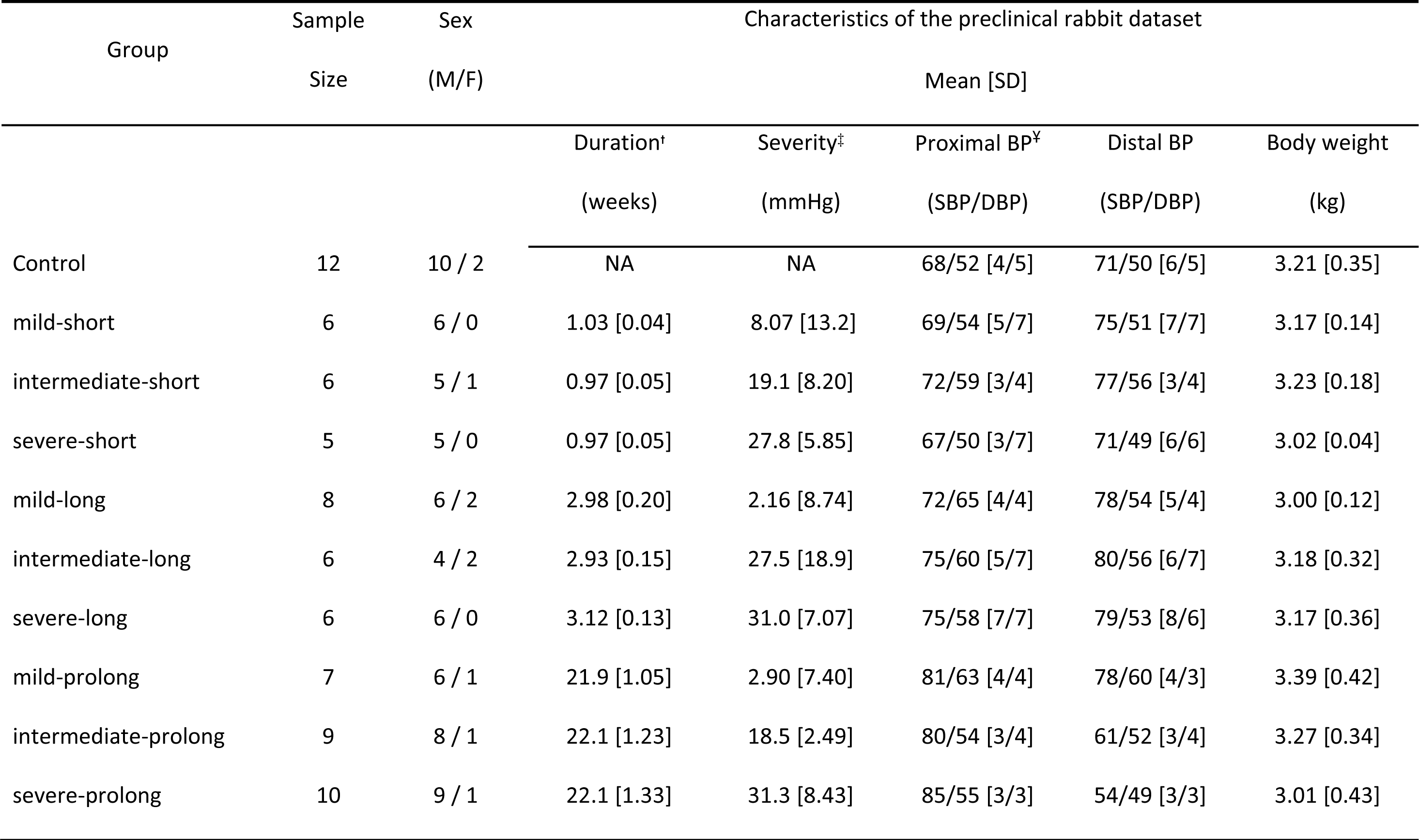
Preclinical rabbit dataset characteristics. ^t^Duration is measured in weeks, long and short CoAs used dissolvable and rapid dissolvable sutures whereas permanent silk sutures were used for prolonged CoA, ^‡^Severity was measured under ∼1 MAC Isoflurane via Doppler-based trans-coarctation peak-to-peak blood pressure gradient [BPGpp; mmHg] in the permanent CoA groups and used to determine the severity experienced by other duration groups prior to the suture dissolving (see Figure S1). BPGpp values have been converted from estimates via simplified Bernoulli equation to catheter representations using: Doppler Gradient = 1.62 × BPGpp + 21.33 from Ghorbannia et al - J Am Soc Echocardiogr. 2022 Dec;35(12):1311-1321. ^¥^Proximal and distal BP were measured under ∼1 MAC Isoflurane after 22 weeks using fluid-filled catheters advanced from carotid and femoral arteries to the aortic arch and descending thoracic aorta (diaphragm level), respectively.

| Group | Sample<br>Size | Sex<br>(M/F) | Characteristics of the preclinical rabbit dataset |  |  |  |  |
| --- | --- | --- | --- | --- | --- | --- | --- |
|  |  |  | Mean [SD] |  |  |  |  |
|  |  |  | Duration<sup>†</sup><br>(weeks) | Severity<sup>‡</sup><br>(mmHg) | Proximal BP<sup>¥</sup><br>(SBP/DBP) | Distal BP<br>(SBP/DBP) | Body weight<br>(kg) |
| Control | 12 | 10 / 2 | NA | NA | 68/52 [4/5] | 71/50 [6/5] | 3.21 [0.35] |
| mild-short | 6 | 6 / 0 | 1.03 [0.04] | 8.07 [13.2] | 69/54 [5/7] | 75/51 [7/7] | 3.17 [0.14] |
| intermediate-short | 6 | 5 / 1 | 0.97 [0.05] | 19.1 [8.20] | 72/59 [3/4] | 77/56 [3/4] | 3.23 [0.18] |
| severe-short | 5 | 5 / 0 | 0.97 [0.05] | 27.8 [5.85] | 67/50 [3/7] | 71/49 [6/6] | 3.02 [0.04] |
| mild-long | 8 | 6 / 2 | 2.98 [0.20] | 2.16 [8.74] | 72/65 [4/4] | 78/54 [5/4] | 3.00 [0.12] |
| intermediate-long | 6 | 4 / 2 | 2.93 [0.15] | 27.5 [18.9] | 75/60 [5/7] | 80/56 [6/7] | 3.18 [0.32] |
| severe-long | 6 | 6 / 0 | 3.12 [0.13] | 31.0 [7.07] | 75/58 [7/7] | 79/53 [8/6] | 3.17 [0.36] |
| mild-prolong | 7 | 6 / 1 | 21.9 [1.05] | 2.90 [7.40] | 81/63 [4/4] | 78/60 [4/3] | 3.39 [0.42] |
| intermediate-prolong | 9 | 8 / 1 | 22.1 [1.23] | 18.5 [2.49] | 80/54 [3/4] | 61/52 [3/4] | 3.27 [0.34] |
| severe-prolong | 10 | 9 / 1 | 22.1 [1.33] | 31.3 [8.43] | 85/55 [3/3] | 54/49 [3/3] | 3.01 [0.43] |

<sup>†</sup>Duration is measured in weeks, long and short CoAs used dissolvable and rapid dissolvable sutures whereas permanent silk sutures were used for prolonged CoA, <sup>‡</sup>Severity was measured under ~1 MAC Isoflurane via Doppler-based trans-coarctation peak-to-peak blood pressure gradient

Machine learning results identified best decision tree regression models to predict thickening, stiffening, impaired contraction, and impaired relaxation. Overall, duration and severity of the CoA were the most important predictors of these active and passive dysfunction indices (see Supplemental Results for normalized importance values and accuracy). Sex had no effect on model prediction in any of the indices.

Figure 2A shows the best decision tree regression fit to aortic thickening patterns observed in rabbits. For mild and short presentation of the CoA, thickness was close to control values. Specifically, data was clustered into 6 leaf nodes with thickness closer to the control group when CoA severity (i.e., BPGpp) was <27.3 mmHg and CoA was present for <20 days. There were two clusters present in this criterion with average thicknesses of 75.27±6.17 and 80.92±6.66 µm/kg, respectively. For the same duration, i.e., <20 days, and BPGpp ≥27.3 mmHg, thickness notably increased to 98.69±13.29 µm/kg. Thickening was more pronounced when CoA was present ≥20 days. Specifically, a thickness of 90.25±7.93 µm/kg was observed when BPGpp<14.3 mmHg and 106.63±10.63 µm/kg when 14.3≤BPGpp<20.0 mmHg. The highest level of thickening (i.e.; 120.55 ±12.14 µm/kg) was observed when BPGpp ≥20.0 mmHg was experienced for ≥20 days.

**Figure 2.** Decision tree regression fit to predict aortic (A) thickening, (B) stiffening, (C) contraction, (D) relaxation in response to CoA-induced mechanical stimuli. For aid in clinical translation, blood pressure gradients (BPGpp) listed have been converted from estimates via simplified Bernoulli equation to catheter representations using the function of Doppler Gradient = 1.62 × BPGpp + 21.33 from Ghorbannia et al - J Am Soc Echocardiogr. 2022;35(12):1311-1321. Thickness values from the descending thoracic aorta distal to the left subclavian and proximal to the suture were quantified from 2D Doppler b-mode image and normalized to body weight [BW; µm/kg]. Stiffness values are in MPa and quantified from aortic segments with hyperelastic material properties characterized via uniaxial extension testing. Active function was assessed by myography for contraction and endothelial-derived relaxation as described in Menon et al - J Pharmacol Toxicol Methods. 2012;65(1):18-28. A portion of the datasets for each index were randomly selected for testing decision tree model performance. Hence, sample sizes are smaller than reported elsewhere in in the study.

Figure 2B shows the best decision tree regression fit to arterial stiffening of rabbit aortic samples proximal to the coarctation. Stiffness remained close to control values for mid and short CoA. Specifically, tree structure was clustered into 4 leaves with stiffness closer to the control group when CoA severity (i.e., BPGpp) was <5.5 mmHg and CoA was present for <20 days. There were two clusters present in this criterion with average stiffnesses of 0.43±0.06 and 0.52±0.06 MPa, respectively. On the other hand, for longer presentations of CoA, duration ≥20 days, stiffening was more pronounced. Specifically, a stiffness of 0.62±0.08 MPa was observed when BPGpp <20.0 mmHg and 0.93±0.23 MPa when BPGpp ≥20.0 mmHg.

Figure 2C shows the best decision tree regression fit to contractile response from rabbit aortic tissue samples proximal to the coarctation. Active contraction response of the aorta remained close to that of control rabbits for mid and short CoAs. Specifically, tree structure was clustered into 6 leaves with contractions distinctively impaired by CoA severity and duration. When severity (i.e., BPGpp) was <2.7 mmHg and CoA was present for <22 days, normalized contraction was close to control (i.e., 3.45±0.29). At the same duration (i.e., <22 days), contraction correlated with body weight when BPGpp ≥2.7 mmHg. Specifically, contraction response decreased to 3.15±0.37 and 2.51±0.30 for body weight < and ≥3.05 kg, respectively. Conversely, for longer presentations of CoA (duration ≥22 days), contraction was more markedly impaired. Specifically, contractile response reduced to 1.94 ±0.32 when BPGpp ≥23.1 mmHg. For BPGpp <23.1 mmHg contractile response decreased to 2.51±0.23 and 2.68±0.23 for durations < and ≥153 days, respectively.

Figure 2D shows the best decision tree regression fit for aortic relaxation observed from CoA rabbits. Active relaxation remained close to that of control rabbits for mid and short CoA. Specifically, tree structure was clustered into 6 leaves with relaxations impaired by CoA severity and duration. When severity (i.e., BPGpp) was <17.2 mmHg and CoA was present for <22 days, normalized relaxation was close to control (i.e., 2.43±0.15 and 2.15±0.21, for durations < and ≥3 days, respectively). Under the same duration (<22 days), normalized relaxation was 1.57±0.59 if exposed to a BPGpp ≥17.2 mmHg. For longer presentations of CoA (duration ≥22 days), relaxation of the aorta was more markedly impaired by severity. Specifically, relaxation reduced to 1.55 ±0.21, 1.07 ±0.27, and 0.79±0.20 for BPGpp <14.3, 14.3 to 23.1 mmHg, and ≥23.1 mmHg, respectively.

### 6.2 Pediatric patients treated for CoA

Descriptive statistics for the CoA patient population are provided in Table 2. Patients with elevated BP are not considered HTN by current guidelines, but 22 of the 239 patients were elevated for systolic BP and 10 for diastolic BP. Thirty-six patients were on at least one antihypertensive medication by their follow-up period, while 174 were not any antihypertensive medication. Medication history was not available for 29 patients.

**Table 2.**
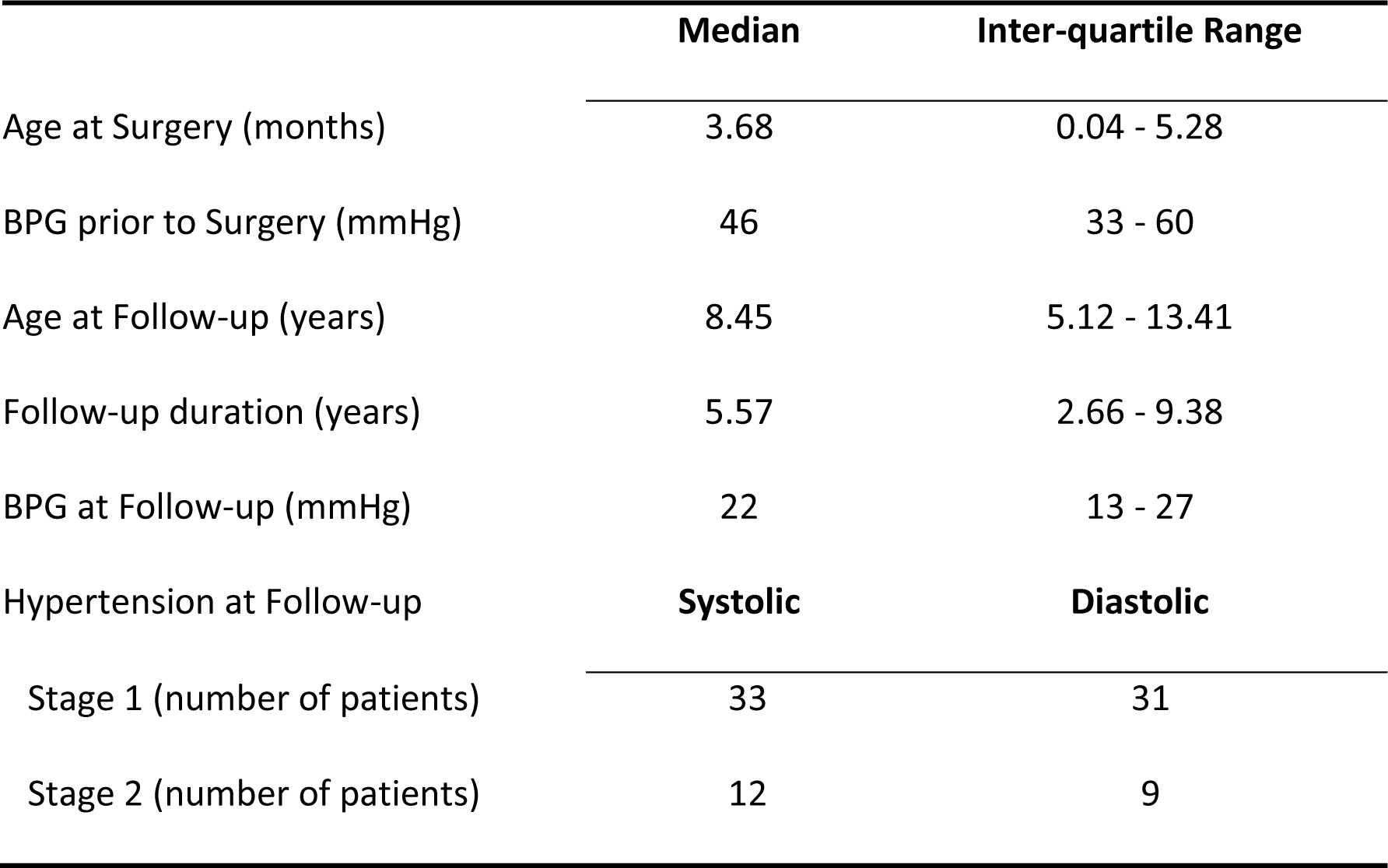
Descriptive statistics for our CoA patient data. BPG=blood pressure gradient estimate by Doppler ultrasound.

|  | Median | Inter-quartile Range |
| --- | --- | --- |
| Age at Surgery (months) | 3.68 | 0.04 - 5.28 |
| BPG prior to Surgery (mmHg) | 46 | 33 - 60 |
| Age at Follow-up (years) | 8.45 | 5.12 - 13.41 |
| Follow-up duration (years) | 5.57 | 2.66 - 9.38 |
| BPG at Follow-up (mmHg) | 22 | 13 - 27 |
| Hypertension at Follow-up | Systolic | Diastolic |
| Stage 1 (number of patients) | 33 | 31 |
| Stage 2 (number of patients) | 12 | 9 |

*Glmulti* produced similar results based on three different information criteria used, where the final model includes two main effects (age at surgery and follow-up time since surgery) as well as four pairwise interactions: (a) age and BPGpp at surgery, (b) age at surgery and body surface area, (c) follow-up time since surgery and BSA, and (d) follow-up time since surgery and hypertensive medication. To avoid incomplete or biased results between predictors and outcome, insignificant main effects associated with significant interactions were also included^23^. The final model is given below with coefficients in Table S1. The receiver operating characteristic (ROC) curve for the model resulting in an AUC of 0.74 is shown in Figure S2.

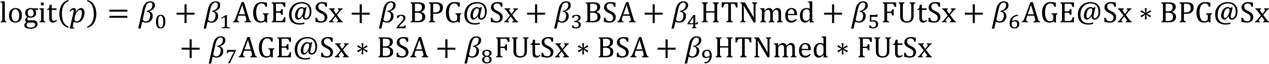

where *p* = likelihood of HTN, AGE@Sx = age at surgery, BPG@Sx = blood pressure gradient at surgery, FUtSx = time since surgery, BSA = body surface area, and HTNmed = hypertensive medication

Similar to preclinical results, decision tree regression (Figure 3) identified both pre-op severity and follow-up time since surgery as main effects with normalized feature importance of 0.56, and 0.25. Interestingly, post-op BPGpp was also observed among main effects with 0.18 normalized feature importance. Specifically, for pre-op BPG <35.7 mmHg, 15% of the patients developed HTN, among which the majority were treated at 3.1 years of age or older and had post-op BPGpp ≥5.9 mmHg. On the other hand, HTN was most prevalent (46%) among patients with pre-op BPGpp ≥35.7 mmHg, among which the majority (72%) were ≥6.4 years old at surgery.

**Figure 3.** Decision tree regression fit to predict hypertension likelihood based on patient datasets. Hypertension status was assessed relative to metrics of duration and severity of the CoA, including, BPGpp and age at surgery (pre-op), as well as the BPGpp and age at follow up (post-op). For aid in clinical translation, blood pressure gradients listed have been converted from estimates via simplified Bernoulli equation to arm-leg pressure gradients (BPGpp) represented using the function of Doppler Gradient = 0.98 × BPGpp + 18.69 from Ghorbannia et al - J Am Soc Echocardiogr. 2022;35(12):1311-1321. A portion of the datasets were randomly selected for testing decision tree model performance. Hence, sample sizes are smaller than reported elsewhere in in the study

Reliability of echo-based measurements revealed ≤8.3% and 10.1% differences among inter- and intra-observer measurements, respectively. The 95% CI for ICC was [0.873, 0.998] for single-subject variability and [0.902, 0.999] on average, which is considered good to excellent reliability^24,25^.

## 7 Discussion

A recent study of 690 patients in the UK with a median surgical repair age of 4 years reported a 70 year survival rate 16% less than the general population with 57% of patients hypertensive at their latest follow-up^26^. This is just one of many studies pointing to persistent morbidity observed since the first surgery for CoA was performed >75 years ago^27^ and underscoring a need for improved treatment guidelines.

There are two main findings from the current work. First, the current BPGpp threshold determined decades ago is likely too high to limit processes leading to persistent aortic changes associated with HTN. Second, severity and duration of the peak-to-peak CoA gradient are important interactive contributors to the likelihood of HTN precursors in the preclinical model as well as HTN status in CoA patients.

While prior work from animal models and patients with CoA reported the importance of coarctation severity relative to the putative treatment BPGpp (i.e., >20 mmHg), the current investigation is the first to systematically evaluate lesser BPGpp severities. BP is similar between species^28^, which provides some basis for the severity ranges from the rabbit model also being applicable to patients with CoA as is discussed more below. Correlation of CoA duration with the degree of structural remodeling and functional impairment could be inferred by prior studies reporting earlier treatment results in less morbidity^18^. The current findings confirm and extend such prior work to elucidate a range of durations that seem to avoid permanent arterial remodeling depending on coarctation severity. The finding of additional predictors being associated with the likelihood of HTN from retrospective data of CoA patients in the current investigation is also consistent with prior studies. For example, in the UK study mentioned above^26^, risk factors for HTN were male sex, older age at repair and/or follow-up as well as residual coarctation.

Figure 3 provides an example of clinical utility from the current work, where the history of the patient (i.e., pre-operative and post-operative BPGpp, age at surgery) can be used to quantify the likelihood of HTN using a simple decision tree framework with these predictors at the leaf nodes. Clinical implementation of these collective predictors allows for a more nuanced and personalized clinical decision-making framework based on coarctation induced mechanical stimuli experienced by each patient. The decision tree will undoubtedly be updated as the current methods are expanded to more patients and additional centers.

There are important shortcomings associated with BPGpp guidelines for CoA that are addressed by the current and ancillary studies. It is difficult to trace the origins of the 20 mmHg and 10 mmHg BPG values in the treatment guidelines. Conversely, the current study provided a systematic approach to evaluating the BPG in CoA patients and using a preclinical model. Having a single BPG cutoff value (e.g. 20 mmHg) regardless of measurement approach (catheterization, arm-leg cuff measurements, or Doppler echocardiography) is concerning. Measurement of the BPGpp is most accurate by catherization, but catheterization is performed on limited basis, usually with stenting or angioplasty. BPGpp can also be estimated via cuffs placed on the arms and legs. However, this is difficult for several reasons (e.g. agitation and/or inappropriately sized cuffs)^29-31^. CoA patients may also have arterial anomalies that limit accurate BPGpp determination^32^. Despite widespread use of the SBE clinically, it has poor agreement with gold-standard catheterization^33^. Collectively these details may explain the suboptimal level of evidence surrounding current treatment guidelines for CoA, and suggests that values in guidelines should depend on the method by which BPGpp is assessed. Related work^11^ has provided further understanding of the relationship between measurement approaches, and establish a new index, the continuous flow pressure gradient (CFPG), which is closely correlated to catheterization and arm-leg cuff BPG data. This index could be used to increase the accuracy of the BPGpp measured and roll out new treatment guidance for CoA patients as part of follow-up research.

Chronic changes in wall tension (the product of radius and BP) exposing arterial cells to pronounced deformation is believed to be the stimuli for arterial thickening. While thickening restores wall stress to a preferred operating range^34^, it also adds stiffness. Decreased strain occurs with age^35^, is a manifestation of arterial remodeling, and is indicative of the risk for HTN^36^. The current study focused on HTN precursors after 22 weeks using the rabbit model of CoA, and prior work by our lab suggests indices of wall shear stress are similar proximally for rabbits exposed to BPG >20 mmHg via permanent or dissolvable sutures at this time point. In contrast, elevated BP, decreased strain, and increased stiffness were localized to the proximal aorta where medial thickening, endothelial dysfunction, and smooth muscle dedifferentiation via myography were present^37^. Importantly, most of the morbidity associated with CoA in patients seems to involve the proximal aorta and its branches. The main findings from the current study are therefore likely a result of adverse mechanical stimuli imposed on the proximal aorta as a result of the coarctation, with changes in BP and deformation primarily implicated (e.g.; strain, wall tension). Recent work^8, 9^ characterized the temporal evolution of these stimuli that ultimately resulted in permanent changes to proximal aortic structure and function^8^ for the groups of rabbits in the current study.

The notion of stimuli proximal to the coarctation influencing adverse remodeling is consistent with prior studies. For example, in dogs, evidence for decreased nitric oxide (NO) bioavailability has been noted above but not below CoA^38^. Multiple studies have reported that the structure and function of the lower limb vasculature of CoA patients is generally normal before and after CoA correction, as opposed to proximal aortic regions, suggesting altered mechanical stimuli from the coarctation are a critical component of long-term morbidity. Kuhn et al^39^ reported impaired distensibility and an index of stiffness in the ascending aorta of neonatal CoA as compared to the descending aorta, and both locations in control patients. While we have not reported distensibility here, the stiffness reported is generally analogous to that in Kuhn et al. Diminished elastic properties of the aorta proximal to the coarctation persisted despite CoA repair in the current study, as in the neonatal CoA patients featured by Kuhn et al. Further prior evidence for impaired function includes an increased response to norepinephrine in the brachial but not a calf artery of CoA vs. control patients^40^, and abnormal brachial but not femoral artery function in repaired CoA patients^41^.

There were several potential study limitations. The results of CoA patient datasets may have been affected by the correlations between different parameters investigated. Age at surgery may also have been affected by other concomitant anomalies like bicuspid aortic valve, long and multi-segment CoA, or other patient complications. Family history was not available for all the patients, limiting assessment of its impact. Adding body mass index (BMI) to the predictors did not improve the model performance, nor did the combination of BMI and BSA. Some values associated with BP categories changed in the midst of the current study (e.g. Stage 2 HTN expressed as >95th percentile + 12 mmHg in prior guidance^14^ applied here vs >99th percentile + 5 mmHg in current guidance^16^). However, Normal and Prehypertension/Elevated definitions have not changed, suggesting that the overall number of HTN CoA patient data sets remains accurate, although the distribution of Stage 1 vs Stage 2 may have shifted slightly if analysis had begun using recent guidelines.

Decreased baroreceptor reflex sensitivity^42^, impaired vascular function^43^, and over activity of the renin-angiotensin system^44^ have all been implicated in the morbidity occurring from CoA (recently reviewed by a portion of the authors^45^). While CoA may be a combination of these issues, the current results show that the duration a coarctation with severity below the current putative treatment threshold is present, in the absence of these other possible contributors, can lead to consequences of proximal aortic stiffening and endothelial dysfunction rooted in pronounced mechanical stimuli^9^.

Association of traditional HTN biomarkers with predictors in the current study has yet to be determined in future work. However, arterial thickening, stiffening, and active arterial response are all determinants of aortic biomechanical behavior previously related to HTN. Since many of these indices can be measured or calculated using clinical imaging modalities, one or more may ultimately serve as important biomarkers for the risk of HTN in CoA once associated normative values are established. Blood pressures measured at the conclusion of the preclinical model duration were lower than expected since rabbits were under ∼1 MAC Isoflurane, but a recent study from our group using computational methods to implement simulated conscious hemodynamic conditions did not observe significant changes in velocity and BPG^11^. Values reported for thickness, contraction and relaxation from the preclinical CoA model were obtained ex vivo and not within an in vivo system where dynamic adjustments to HTN are being made.

It is difficult to directly conclude that results concerning severity and duration of CoA in the rabbit directly agree with those from the CoA patients. Nonetheless, BPGpp severity and at surgery being the main factors of the decision tree to predict HTN suggests that findings from the rabbit model may translate to patients with CoA. Applying our findings as simply as possible shows that endothelial function is maintained in response to a BPGpp <13 mmHg when present for <3 weeks using our preclinical model. Plotting the age of New Zealand White rabbits^10^ versus that of patients suggests there may be a ∼1.5 year window of aortic plasticity for treating patients with CoA diagnosed around age 9 (Figure 4). Knowing that arterial stiffening occurs with age, this window is likely longer if patients are treated sooner, and possibly shorter if diagnosis occurs later. This hypothesis regarding a window of plasticity based on our rabbit CoA results remains to be directly tested in future studies, along with whether revised guidelines using less severe BPGpp and careful assessment of CoA duration are possible to manage clinically. Adapting to new evidence may also take some time since the morphology of a lesser BPGpp may not necessarily appear severe.

**Figure 4.** Human age plotted as a function of New Zealand White rabbit age. Rabbits undergo CoA at 10 weeks of age. One of the results presented shows endothelial function via myography is maintained in response to a BPGpp <13 mmHg if present for <3 weeks in our preclinical CoA model. Extracting the corresponding human age from the plot suggests there may be a ∼1.5 year window of aortic plasticity for treatment in children with CoA manifesting around age 9 years old.

## 8 Conclusions

The current findings emphasize the importance of both severity and duration of the peak-to-peak CoA gradient to the risk of HTN precursors in the preclinical rabbit model as well as HTN status in patients. These findings provide a foundation for revising CoA treatment guidelines to limit the likelihood of HTN.

## 9 Perspectives

This study used a preclinical model to systematically evaluate blood pressure gradients observed clinically from aortic coarctation, with relevance to CoA patients tested by retrospective review of predictors (pre-operative blood pressure gradient, surgical age, etc.) versus follow-up hypertensive status. Results show current blood pressure gradient thresholds are likely too high to prevent aortic changes associated with hypertension, and that the severity and duration of the peak-to-peak blood pressure gradient across the coarctation should both be considered to decrease the risk of hypertension in pediatric aortic coarctation patients. We are optimistic that the current study will serve a basis for revisiting current guidelines for treatment of aortic coarctation based on the blood pressure gradient.

## Data Availability

All data produced in the present study are available upon reasonable request to the authors.

## 3 Abbreviations and acronyms

ACh: Acetylcholine
AUC: Area under the receiver operative curve
AUD: Area under the dose response curve
BMI: body mass index
BP: Blood Pressure
BPG: Blood Pressure Gradient
BPGpp: Peak-to-peak blood pressure gradient
BSA: Body surface area
BW: Body weight
CFPG: continuous flow pressure gradient
CoA: Coarctation of the Aorta
HTN: Hypertension
ICC: intraclass correlation coefficient
PE: Phenylephrine
SM: Smooth muscle

## 10 Acknowledgements

a. Acknowledgements: The authors thank Angie Klemm, Alexander Armstrong, Ken Allen, Eric Jensen, Brandon Wegter, Angelia Espinal, Nick Peterson, and Andrew Spearman for their assistance at various points throughout this study.
b. Sources of Funding: Research reported was partially supported by the American Heart Association under award number 15GRNT25700042 (JFL) as well as the National Institutes of Health - National Heart, Lung, and Blood Institute under award numbers R01HL142955 (JFL) and R15HL096096 (JFL). The content is solely the responsibility of the authors and does not necessarily represent the official views of the National Institutes of Health or the American Heart Association.
c. Disclosure: none

## 12 Novelty and Relevance

### What Is New?

A machine learning based decision tree analysis revealed that predictors including pre-operative blood pressure gradient, post-operative residual blood pressure gradient, and surgical age statistically correlate with the risk of hypertension in treated aortic coarctation patients. This machine learning based decision tree approach allows for straightforward assessment of the risk for hypertension after treatment for aortic coarctation using patient-specific predictors.

### What is Relevant?

For unknown reasons, treated coarctation of the aorta patients often have a reduced life expectancy from cardiovascular morbidity, primarily hypertension. The blood pressure gradient induced by the coarctation is a primary indication for treatment, but there is a lack of data for the current >20 mmHg value. This study systematically identified specific values for blood pressure gradient severity and the duration it is present to avoid persistent arterial remodeling and endothelial dysfunction in a preclinical model, and related these predictors to hypertensive status in pediatric patients treated for coarctation of the aorta. These findings provide a foundation for revising guidelines to limit the risk of hypertension after treatment for coarctation of the aorta.

### Clinical/Pathologic Implications?

Our study suggests current blood pressure based thresholds related to coarctation of the aorta are likely too high to prevent aortic changes associated with hypertension, and that the severity and duration of the peak-to-peak blood pressure gradient across a coarctation should both be considered to decrease the risk of hypertension in these patients. Examples of a machine learning based decision tree approach that uses retrospective data from coarctation patients have been provided as a starting point for revisiting treatment guidelines.

## 11 References

1. Roger VL, Go AS, Lloyd-Jones DM, Adams RJ, Berry JD, Brown TM, Carnethon MR, Dai S, de Simone G, Ford ES, Fox CS, Fullerton HJ, Gillespie C, Greenlund KJ, Hailpern SM, Heit JA, Ho PM, Howard VJ, Kissela BM, Kittner SJ, Lackland DT, Lichtman JH, Lisabeth LD, Makuc DM, Marcus GM, Marelli A, Matchar DB, McDermott MM, Meigs JB, Moy CS, Mozaffarian D, Mussolino ME, Nichol G, Paynter NP, Rosamond WD, Sorlie PD, Stafford RS, Turan TN, Turner MB, Wong ND and Wylie-Rosett J. Heart disease and stroke statistics--2011 update: a report from the American Heart Association. Circulation. 2011;123:e18–e209.

2. Cohen M, Fuster V, Steele PM, Driscoll D and McGoon DC. Coarctation of the aorta. Long-term follow-up and prediction of outcome after surgical correction. Circulation. 1989;80:840–5.

3. Isselbacher EM, Preventza O, Hamilton Black J, 3rd, Augoustides JG, Beck AW, Bolen MA, Braverman AC, Bray BE, Brown-Zimmerman MM, Chen EP, Collins TJ, DeAnda A, Jr., Fanola CL, Girardi LN, Hicks CW, Hui DS, Schuyler Jones W, Kalahasti V, Kim KM, Milewicz DM, Oderich GS, Ogbechie L, Promes SB, Gyang Ross E, Schermerhorn ML, Singleton Times S, Tseng EE, Wang GJ and Woo YJ. 2022 ACC/AHA Guideline for the Diagnosis and Management of Aortic Disease: A Report of the American Heart Association/American College of Cardiology Joint Committee on Clinical Practice Guidelines. Circulation. 2022;146:e334-e482.

4. Torok RD, Campbell MJ, Fleming GA and Hill KD. Coarctation of the aorta: Management from infancy to adulthood. World J Cardiol. 2015;7:765–75.

5. Warnes CA, Williams RG, Bashore TM, Child JS, Connolly HM, Dearani JA, Del Nido P, Fasules JW, Graham TP, Jr., Hijazi ZM, Hunt SA, King ME, Landzberg MJ, Miner PD, Radford MJ, Walsh EP and Webb GD. ACC/AHA 2008 Guidelines for the Management of Adults with Congenital Heart Disease: Executive Summary: a report of the American College of Cardiology/American Heart Association Task Force on Practice Guidelines (writing committee to develop guidelines for the management of adults with congenital heart disease). Circulation. 2008;118:2395–451.

6. Burns PB, Rohrich Rj Fau - Chung KC and Chung KC. The levels of evidence and their role in evidence-based medicine. Plast Reconstr Surg. 2011;128:305–10.

7. Rosenthal E. Stent implantation for aortic coarctation: the treatment of choice in adults? J Am Coll of Cardiol. 2001;38:1524–27.

8. Ghorbannia A, Maadooliat M, Woods RK, Audi SH, Tefft BJ, Chiastra C, Ibrahim EH and LaDisa JF, Jr. Aortic remodeling kinetics in response to coarctation-induced mechanical perturbations. Biomedicines. 2023;11:1817.

9. Azarnoosh J, Ghorbannia A, Ibrahim EH, Jurkiewicz H, Kalvin L and LaDisa JF, Jr. Temporal evolution of mechanical stimuli from vascular remodeling in response to the severity and duration of aortic coarctation using a preclinical model. Nature: Scientific Reports. 2023;13:8352.

10. Dutta S and Sengupta P. Rabbits and men: relating their ages. J Basic Clin Physiol Pharmacol. 2018;29:427–435.

11. Ghorbannia A, Ellepola CD, Woods RK, Ibrahim EH, Maadooliat M, Ramirez HM and LaDisa JF, Jr. Clinical, Experimental, and Computational Validation of a New Doppler-Based Index for Coarctation Severity Assessment. J Am Soc Echocardiogr. 2022;35:1311–1321.

12. Wendell DC, Friehs I, Samyn MM, Harmann LM and LaDisa JF, Jr. Treating a 20 mm Hg gradient alleviates myocardial hypertrophy in experimental aortic coarctation. The Journal of surgical research. 2017;218:194–201.

13. Menon A, Wendell DC, Wang H, Eddinger TJ, Toth JM, Dholakia RJ, Larsen PM, Jensen ES and LaDisa JF, Jr. A coupled experimental and computational approach to quantify deleterious hemodynamics, vascular alterations, and mechanisms of long-term morbidity in response to aortic coarctation. J Pharmacol Toxicol Methods. 2012;65:18–28.

14. Flynn JT, Kaelber DC, Baker-Smith CM, Blowey D, Carroll AE, Daniels SR, de Ferranti SD, Dionne JM, Falkner B, Flinn SK, Gidding SS, Goodwin C, Leu MG, Powers ME, Rea C, Samuels J, Simasek M, Thaker VV, Urbina EM, Subcommittee On Screening and Management Of High Blood Pressure In Children. Clinical Practice Guideline for Screening and Management of High Blood Pressure in Children and Adolescents. Pediatrics. 2017;140. Erratum: 2017; 140(3):e20171904

15. Rosner B, Cook N, Portman R, Daniels S and Falkner B. Determination of blood pressure percentiles in normal-weight children: Some methodological issues. American Journal of Epidemiology. 2008;167:653–666.

16. National High Blood Pressure Education Program Working Group on High Blood Pressure in Children and Adolescents. The fourth report on the diagnosis, evaluation, and treatment of high blood pressure in children and adolescents. Pediatrics. 2004;114:555-76.

17. Shypailo RJ. Age-based Pediatric Blood Pressure Reference Charts. 2018. Baylor College of Medicine, Children’s Nutrition Research Center, Body Composition Laboratory Web https://www.bcm.edu/bodycomplab/BPappZjs/BPvAgeAPPz.html

18. Panzer J, Bove T, Vandekerckhove K and De Wolf D. Hypertension after coarctation repair-a systematic review. Transl Pediatr. 2022;11:270–279.

19. Stoica P and Selen Y. Model-order selection: a review of information criterion rules. IEEE Signal Processing Magazine. 2004;21:36–47.

20. Menard S. Six approaches to calculating standardized logistic regression coefficients. The American Statistician. 2004;58:218–223.

21. Calcagno V and de Mazancourt C. glmulti: An R Package for Easy Automated Model Selection with (Generalized) Linear Models. Journal of Statistical Software. 2010;34:1-29.

22. Pedregosa F, Varoquaux G, Gramfort A, Michel V, Thirion B, Grisel O, Blondel M, Prettenhofer P, Weiss R, Dubourg V, Vanderplas J, Passos A, Cournapeau D, Brucher M, Perrot M and Duchesnay E. Scikit-learn: Machine Learning in Python. J Mach Learn Res. 2011;12:2825–2830.

23. Aiken LS and West SG. Multiple regression: Testing and interpreting interaction: New York: Sage Publications, Inc; 1991.

24. Cicchetti DV. Guidelines, criteria, and rules of thumb for evaluating normed and standardized assessment instruments in psychology. Psychological Assessment. 1994;6:284–290.

25. Koo TK and Li MY. A Guideline of Selecting and Reporting Intraclass Correlation Coefficients for Reliability Research. J Chiropr Med. 2016;15:155–63. Erratum: 2017; 16(4): 346.

26. Lee MGY, Babu-Narayan SV, Kempny A, Uebing A, Montanaro C, Shore DF, d’Udekem Y and Gatzoulis MA. Long-term mortality and cardiovascular burden for adult survivors of coarctation of the aorta. Heart. 2019;105:1190–1196.

27. Kvitting JP and Olin CL. Clarence Crafoord: a giant in cardiothoracic surgery, the first to repair aortic coarctation. Ann Thorac Surg. 2009;87:342–6.

28. Wolinsky H and Glagov S. A lamellar unit of aortic medial structure and function in mammals. Circ Res. 1967;20:99–111.

29. Alvarez J, Aguilar F and Lurbe E. Blood pressure measurement in children and adolescents: key element in the evaluation of arterial hypertension. An Pediatr (Engl Ed). 2022;96:536 e1–536 e7.

30. Palatini P and Asmar R. Cuff challenges in blood pressure measurement. J Clin Hypertens (Greenwich). 2018;20:1100–1103.

31. Bird C and Michie C. Measuring blood pressure in children. BMJ. 2008;336:1321.

32. Hoffman JI. The challenge in diagnosing coarctation of the aorta. Cardiovasc J Afr. 2018;29:252–255.

33. Feltes TF, Bacha E, Beekman RH, 3rd, Cheatham JP, Feinstein JA, Gomes AS, Hijazi ZM, Ing FF, de Moor M, Morrow WR, Mullins CE, Taubert KA and Zahn EM. Indications for cardiac catheterization and intervention in pediatric cardiac disease: a scientific statement from the American Heart Association. Circulation. 2011;123:2607–52.

34. Wolinsky H and Glagov S. Comparison of abdominal and thoracic aortic medial structure in mammals. Deviation of man from the usual pattern. Circ Res. 1969;25:677–86.

35. Morrison TM, Choi G, Zarins CK and Taylor CA. Circumferential and longitudinal cyclic strain of the human thoracic aorta: age-related changes. J Vasc Surg. 2009;49:1029–36.

36. Liu SQ and Fung YC. Relationship between hypertension, hypertrophy, and opening angle of zero-stress state of arteries following aortic constriction. J Biomech Eng. 1989;111:325–35.

37. Menon A, Eddinger TJ, Wang H, Wendell DC, Toth JM and LaDisa JF, Jr. Altered hemodynamics, endothelial function, and protein expression occur with aortic coarctation and persist after repair. American journal of physiology Heart and circulatory physiology. 2012;303:H1304–18.

38. Barton CH, Ni ZM and Vaziri ND. Enhanced nitric oxide inactivation in aortic coarctation-induced hypertension. Kidney Int. 2001;60:1083–1087.

39. Kuhn A, Baumgartner D, Baumgartner C, Horer J, Schreiber C, Hess J and Vogt M. Impaired elastic properties of the ascending aorta persist within the first 3 years after neonatal coarctation repair. Pediatr Cardiol. 2009;30:46–51.

40. Gidding SS, Rocchini AP, Moorehead C, Schork MA and Rosenthal A. Increased forearm vascular reactivity in patients with hypertension after repair of coarctation. Circulation. 1985;71:495–9.

41. Gardiner HM, Celermajer DS, Sorensen KE, Georgakopoulos D, Robinson J, Thomas O and Deanfield JE. Arterial reactivity is significantly impaired in normotensive young adults after successful repair of aortic coarctation in childhood. Circulation. 1994;89:1745–50.

42. Polson JW, McCallion N, Waki H, Thorne G, Tooley MA, Paton JF and Wolf AR. Evidence for cardiovascular autonomic dysfunction in neonates with coarctation of the aorta. Circulation. 2006;113:2844–50.

43. Brili S, Tousoulis D, Antoniades C, Aggeli C, Roubelakis A, Papathanasiu S and Stefanadis C. Evidence of vascular dysfunction in young patients with successfully repaired coarctation of aorta. Atherosclerosis. 2005;182:97–103.

44. Parker FB, Jr., Streeten DH, Farrell B, Blackman MS, Sondheimer HM and Anderson GH, Jr. Preoperative and postoperative renin levels in coarctation of the aorta. Circulation. 1982;66:513–4.

45. Kenny D and LaDisa JF, Jr. Aortic coarctation: clinical concepts, engineering applications and impact of an integrated medico-engineering approach.. In: G. Butera, Schievano, S., Biglino, G., McElhinney, D.B., ed. Modelling Congenital Heart Disease: Springer; 2022.

