## Appendix for "Coarctation duration and severity predict risk of hypertension precursors in a preclinical model and hypertensive status among patients"

**Short Title**: Duration and severity of CoA predict hypertension

**Conflict of interest**: The authors declare that the research was conducted in the absence of any commercial or financial relationships that could be construed as a potential conflict of interest

**Supplemental Methods**

***Preclinical model of CoA***

Experimental procedures were approved by applicable Animal Care and Use Committees. All procedures conformed to the National Institutes of Health Guide for the Care and Use of Laboratory Animals. After >72 hours of acclimation upon receipt from Kuiper Rabbit Ranch, New Zealand white rabbits ~10 weeks old and weighing ~1.0 kg were anesthetized with ketamine/xylazine IM and maintained with isoflurane (minimum alveolar concentration ≈ 1%)[1]. Rabbits (total 75; N=5-12/group) were randomly designated (simple randomization) to surgically undergo discrete CoA of varying severity and associated mechanical stimuli via left thoracotomy in the third intercostal space by tying suture around the aorta against a wire of known diameter (1.6, 2.0 or 2.7 mm). Removal of the wire resulted in CoA severity dependent on the wire diameter used and resulting in BPGpp within the range observed clinically (mild: BPGpp ≤12 mmHg, intermediate: BPGpp 13 - 19 mmHg, and severe: BPGpp ≥20 mmHg). Sutures were tied around the proximal descending thoracic aorta (distal to the left subclavian branch) where CoA most often presents clinically. To investigate the effect of duration of the mechanical stimuli caused by CoA, sutures with different dissolving properties (i.e., rapid dissolvable Vicryl, dissolvable Vicryl, or permanent silk) were used to initiate the stenosis for different durations (short: ~1 weeks, long: 3 weeks, and prolonged: 22 weeks) at the severity levels mentioned. This resulted in 9 experimental groups (3 severities and 3 durations) as well as a group of non-experimental control littermate rabbits. Subcutaneous Furosemide (1-5 mg/kg) injections were administered as needed for up to 14 days post-surgery to limit and/or treat edema. Rabbits were housed in Allentown rack cage systems. Pans beneath each cage are filled with P.J. Murphy Sani-Chips. Rabbits are fed LabDiet 5326 that is supplemented with Timothy hay and apple slices approximately twice per week. Additional experimental and postprocedural monitoring details generally applied with animals in the current study can be found in our prior work[2]. The number of rabbits used for this study represents the minimum to meet the statistical objectives of the study, consistent with the principles of replacement, reduction, and refinement.

***CoA severity classification in the preclinical model based on evolution of the Doppler gradient***

Weekly Doppler ultrasound imaging was performed by a trained sonographer using protocols similar to human echocardiography[3] until BPGpp estimates via of the BP gradient across the CoA calculated using the simplified Bernoulli Equation no longer changed with body weight. For dissolvable CoA groups, BP was expected to recover to normal as the suture dissolved. Severity for rabbits in these groups was estimated noninvasively using Doppler BPGpp evolution curves compared to those from the permanent CoA groups. Specifically, the temporal evolution of Doppler BPG and BPGpp via catheter measurements in the permanent CoA group (22-week duration) were used to determine the severity of coarctations experienced by rabbits in the other duration groups prior to dissolving of the suture (Figure 1S). Rabbits were re-anaesthetized after ~22 weeks for catheter-based measurement of simultaneous BP waveforms proximal and distal to the CoA[2].

***Assessing precursors of hypertension (HTN) using the preclinical rabbit model of CoA***

Aortic active and passive response was assessed relative to the severity and duration of the CoA generated using experimental protocols explained below. Indices of aortic remodeling including thickness, stiffness, and impaired vasoactive response by myography were used as surrogates for HTN. Detailed results are noted elsewhere[4, 5] with an aggregate measure of alterations (i.e., area under the dose-response and hyperelastic curves) and multiple linear regression fits presented in the manuscript.

*Thickness of the aorta*

Doppler b-mode images were used to quantify wall thickness in the proximal descending thoracic aorta from coarctation-induced mechanical stimuli[5]. Measurements were made in triplicate with the mean value reported for each date. Body weight was also measured weekly using a scientific scale and the thickness evolution was normalized to interpolated body weight.

*Stiffness of the aorta*

Aortas also underwent material characterization by uniaxial extension testing (MTS Criterion Load Frame, MTS, Minneapolis, U.S.A.) at 37$^{O}$C in an environmental chamber (MTS Bionix EnviroBath, Minneapolis, U.S.A.). Samples were dissected with a length-to-width ratio of ~ 2.6 and preconditioned by stretching to 10% of the gauge length. Extension was performed at 10 mm/min until hyperelastic behavior was observed. Resulting stress-stretch curves were used to characterize stiffness. Area under the stress-stretch curve (i.e., strain energy in the sample) was quantified in the stretch range of 1 to 2 as an aggregate measure of overall stiffness.

*Functional response of the aorta*

Functional changes were quantified by wire myography of aortic segments (3-4 mm rings). Briefly, active function testing was conducted to observe smooth muscle (SM) contraction via phenylephrine (PE) as well as endothelial-dependent relaxation by acetylcholine (ACh) in a half-log increasing dose response from $10^{-9}$ to $10^{-5}$ M concentrations. Arteries were pre-contracted with PE to the half-maximal effective concentration (EC50) and cumulative addition of agonists was initiated to plateau. Contractile and relaxation response curves were quantified as a percentage of precontracted active tension. Area under the dose response curves (AUD) was then quantified as an aggregate measure of dysfunction. Quantifications were performed in duplicate for paired myography channels from the proximal descending thoracic aorta.

**HTN in pediatric patients treated for CoA**

Electronic medical records of treated CoA patients (N=239) were retrospectively studied after exempt determination by the Institutional Review Boards at the Medical College of Wisconsin and Shand's Children's Hospital. Briefly, BPGpp, patient age, sex, height, weight, medication history, and follow-up BP were used to determine BP percentiles and HTN status. Patients <18 years at follow-up were assessed using the Clinical Practice Guideline for Screening and Management of High Blood Pressure in Children and Adolescent[6-8], which was implemented via an online calculator from Baylor College of Medicine[9]. Patients ≥18 years at follow-up were assessed using Joint National Committee 7 guidelines for systolic/diastolic BP: normal (<120/80 mmHg), pre-HTN (120-139/80-89 mmHg), HTN (includes Stage 1: 140-159/90-99 mmHg and Stage 2: >160/>100 mmHg). Patients with BP percentiles registering as Stage 1 or Stage 2 for systolic or diastolic BP were interpreted as hypertensive. BPGpp in patient records was determined from echocardiographic Doppler-based estimates of peak velocity by applying the simplified Bernoulli equation (SBE). BPGpp noted as "not significant" or "none" were interpreted as zero and average values were used when a range was noted.

**Statistical Analysis**

Multivariate regression analysis was performed on pre-clinical rabbit datasets with severity, duration, body weight, and sex as predictors. Predictors were considered relative to aspects of CoA that could impact aortic remodeling and HTN in a preclinical model, as well as corresponding indices from retrospective data and systematic review of HTN prevalence in CoA patients[5, 10]. Forward and backward stepwise selection methods were used to ensure the best model was not over-/under-fitting. Three different information criteria, i.e., Akaike information criteria (AIC), corrected Akaike information criteria (AICc), and Bayesian information criteria (BIC), were used to identify the best model[11]. Normality of the residuals were tested through D'Agostino-Pearson, Anderson-Darling, Shapiro-Wilk, and Kolmogorov-Smirnov tests. Numerical predictors were scaled by dividing by the corresponding standard deviation so as to make these predictors scale-free and improve comparability of variables[12].

Similarly, to assess the possibility of translating any new BPGpp threshold ranges to patients, predictors from hypertensive CoA patients at follow-up were analyzed from 260 million possible models (e.g., logistic regression, support vector machine, decision trees, etc.). Predictors included pre & post-operative estimates of BPGpp (i.e., severity), surgery date versus date of birth (i.e., duration), follow-up time since surgery, sex, body surface area (BSA) and knowledge of anti-hypertensive medication (where available). Possible models also included the respective 21 pairwise interactions of these seven predictors. The best model was found with *Glmulti*, an R package utilizing a genetic algorithm[13].

An interpretable machine learning algorithm, the Classification and Regression Tree, was additionally used to model the impact of the predictors on HTN precursors in the preclinical model, and HTN status from treated CoA patient data sets. *Sklearn*, a classification learner package[14], was used for this purpose. Model predictions were then used to compute predictive performance metrics for each approach.

Reliability of echo-based measurements was investigated through intraclass correlation coefficient (ICC) using SPSS software. Two observers obtained quantities for a random subgroup of rabbit and patient data (n=10).

### Supplemental Results

#### Preclinical model of CoA - normalized importance values and accuracy for predictors contributing to precursors of hypertension

Machine learning results identified best decision tree regression models to predict thickening, stiffening, impaired contraction, and impaired relaxation. Overall, duration and severity of the CoA were the most important predictors of these active and passive dysfunction indices. The normalized importance of duration was 0.57, 0.58, 0.59, and 0.53 for arterial thickening, stiffening, contraction impairment, and relaxation impairment, respectively. The normalized importance of severity was 0.43, 0.43, 0.33, and 0.35 for arterial thickening, stiffening, contraction impairment, and relaxation impairment, respectively. On the other hand, body weight had only a minor effect on model predictions with normalized importance values of 0.00, 0.00, 0.08, 0.12 for arterial thickening, stiffening, contraction impairment, and relaxation impairment, respectively. Sex had no effect on model prediction in any of the metrics of dysfunction studied.

Model performance for the thickness decision tree regression fit was assessed for an assumed threshold equal to higher $75^{th}$, $90^{th}$, and $99^{th}$ quantile of the thicknesses in the control group, i.e., 83.5, 84.1, and 91.2, respectively. The corresponding accuracy, sensitivity, and specificity were [0.74, 0.00, 1.00], [0.79, 0.80, 0.79], and [0.79, 0.67, 0.90], respectively.

Model performance for the stiffening decision tree regression fit was assessed for an assumed threshold equal to higher $75^{th}$, $90^{th}$, and $99^{th}$ quantile of the stiffnesses observed in the control group, i.e., 0.323, 0.350, 0.365. The corresponding accuracy, sensitivity, and specificity were [0.74, 0.00, 1.00], [0.79, 0.80, 0.79], and [0.79, 0.67, 0.90], respectively.

Model performance for the contraction decision tree regression was assessed for an assumed threshold equal to lower $1^{st}$, $10^{th}$, and $25^{th}$ quantile of the contraction observed in the control group, i.e., 2.91, 3.17, and 3.23. The corresponding[accuracy, sensitivity, and specificity were [0.79, 0.82, 0.75], [0.89, 1.00, 0.71], and [0.89, 0.93, 0.80], respectively.

Model performance for the relaxation decision tree regression fit was assessed for an assumed threshold equal to lower $1^{st}$, $10^{th}$, and $25^{th}$ quantile of the contraction observed in the control group, i.e., 2.128, 2.200, 2.29. The corresponding accuracy, sensitivity, specificity were [0.68, 0.62, 0.83], [0.84, 0.92, 0.67], and [0.84, 0.87, 0.75], respectively.

#### Pediatric patients treated for CoA

The final model from multivariate regression model is provided in the main manuscript with coefficients in Table S1. The receiver operating characteristic (ROC) curve for the model described above resulting in an AUC of 0.74 is shown in Figure S2.

### Supplemental Table

Table S1. Coefficients for Likelihood of Hypertension using the Logistic Regression Model.

$$\mathrm{logit}(p) = \beta_0 + \beta_1\mathrm{AGE@Sx} + \beta_2\mathrm{BPG@Sx} + \beta_3\mathrm{BSA} + \beta_4\mathrm{HTNmed} + \beta_5\mathrm{FUtSx} + \beta_6\mathrm{AGE@Sx} * \mathrm{BPG@Sx} + \beta_7\mathrm{Age@Sx} * \mathrm{BSA} + \beta_8\mathrm{FUtSx} * \mathrm{BSA} + \beta_9\mathrm{HTNmed} * \mathrm{FUtSx}$$

where $p$ = likelihood of HTN, AGE@Sx = age at surgery, BPG@Sx = blood pressure gradient at surgery, FUtSx = time since surgery, BSA = body surface area, and HTNmed = hypertensive medication, AIC=226.4

| Terms | Coefficient Metrics | | | | | | | Odds Ratio (OR) Metrics | | | | |
|---|---|---|---|---|---|---|---|---|---|---|---|---|
| | p-values | Coeff.‡ | SD | SD Interval | | 95% CI | | OR | SD Interval | | 95% CI | |
| | | | | 1-SD | 1+SD | Lower | Upper | | 1-SD | 1+SD | Lower | Upper |
| Intercept | 0.393 | 0.603 | 0.706 | -0.102 | 1.309 | -0.780 | 1.986 | 1.828 | 0.903 | 3.702 | 0.459 | 7.289 |
| Age at Surgery (Age@Sx) | 0.003** | -5.64 | 1.866 | -7.506 | -3.774 | -9.297 | -1.982 | 0.004 | 0.001 | 0.023 | 0.000 | 0.138 |
| Pre-op BPGpp (BPG@Sx) | 0.451 | 0.149 | 0.197 | -0.048 | 0.346 | -0.238 | 0.535 | 1.160 | 0.953 | 1.414 | 0.788 | 1.708 |
| Body surface area (BSA) | 0.487 | -0.381 | 0.547 | -0.928 | 0.167 | -1.453 | 0.692 | 0.684 | 0.395 | 1.181 | 0.234 | 1.997 |
| Medication (HTNmed) | 0.876 | -0.115 | 0.735 | -0.850 | 0.620 | -1.555 | 1.325 | 0.891 | 0.428 | 1.859 | 0.211 | 3.764 |
| Time since Surgery (FUtSx) | 0.001** | -2.161 | 0.669 | -2.831 | -1.492 | -3.473 | -0.850 | 0.115 | 0.059 | 0.225 | 0.031 | 0.428 |
| Age@Sx: BPG@Sx | 0.030* | 0.883 | 0.406 | 0.477 | 1.289 | 0.087 | 1.679 | 2.418 | 1.611 | 3.631 | 1.090 | 5.363 |
| Age@Sx:BSA | 0.005** | 0.937 | 0.333 | 0.604 | 1.270 | 0.285 | 1.590 | 2.553 | 1.830 | 3.562 | 1.329 | 4.904 |
| FUtSx:BSA: | 0.009** | 0.521 | 0.200 | 0.320 | 0.721 | 0.128 | 0.913 | 1.683 | 1.377 | 2.056 | 1.136 | 2.493 |
| HTNmed:FUtSx | 0.073• | 1.208 | 0.673 | 0.535 | 1.881 | -0.110 | 2.526 | 3.347 | 1.708 | 6.557 | 0.896 | 12.507 |

‡Standardization was performed to improve interpretation of the effect sizes, i.e., OR metrics. Continuous variables (Age@Sx, BPG@Sx, FUtSx, and BSA) were scaled by dividing by SD, therefore, removing the dimensionality.

CI = 95% confidence interval, OR: odds ratio, SD = standard deviation.

Significance codes: 0 ‘ *** ’ 0.001 ‘ ** ’ 0.01 ‘ * ’ 0.05 ‘ • ’ 0.1 ‘ ’ 1

Major Resources Table

**Animals (in vivo studies)**

| Species | Vendor or Source | Background Strain | Sex | Persistent ID / URL |
|---|---|---|---|---|
| Oryctolagus cuniculus | Kuiper Rabbit Ranch | New Zealand white | M/F | N/A |

**Supplemental Figures**

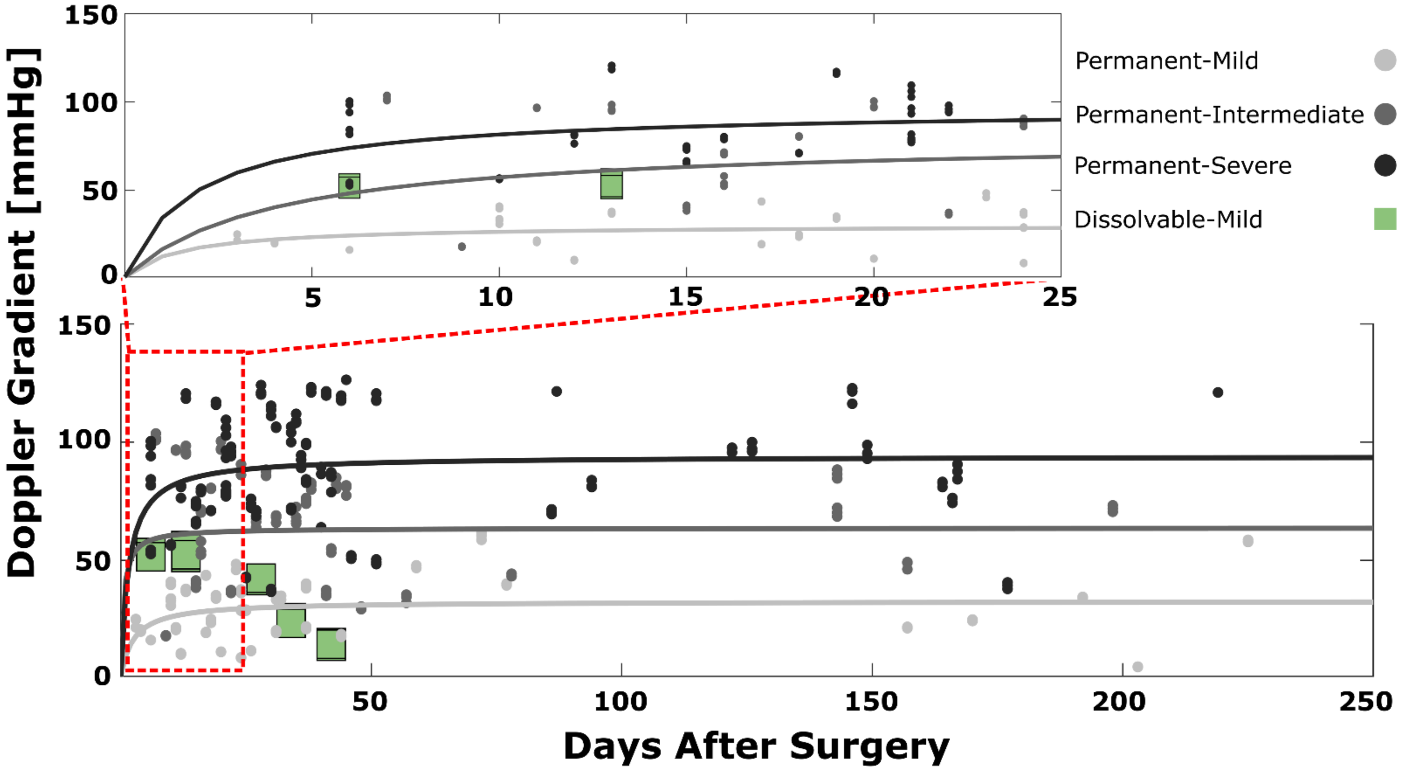


*Figure S1. CoA severity classification based on evolution of the Doppler gradient. For permanent sutures, Doppler gradients stabilized at specific severities determined by the wire diameter used to create the coarctation (shades of gray indicating different severity groups). The highlighted example (green boxes) is from a rabbit representative of the cohort with intermediate CoA severity (i.e., BPGpp 13 - 20 mmHg) before suture absorption (~3-week period, red inset). Green boxes depict initial increases in Doppler gradients following introduction of the dissolvable suture used to create CoA, which is followed by a decrease as the sutures dissolved. The Doppler gradient calculation used peak jet velocity readings in a simplified Bernoulli equation modified to correct for proximal acceleration. In the permanent CoA group, gradients were compared to invasive BPGpp measured at the end of the experimental duration using fluid-filled catheters. Conversely, for dissolvable sutures, only non-invasive ultrasound measurements were used because blood pressure recovered by the end of the protocol. Leveraging the Doppler BPG measurements in the permanent CoA cohort, BPGpp were interpreted for assignment into dissolvable groups according to their peak Doppler-based measurements. BPGpp listed have been converted from estimates via simplified Bernoulli equation to catheter representations using the function of Doppler Gradient = 1.62 × BPGpp + 21.33 from Ghorbannia et al - J Am Soc Echocardiogr. 2022 Dec;35(12):1311-1321. BP: blood pressure, BPG: blood pressure gradient, BPGpp: peak-to-peak blood pressure gradient across the CoA, CoA: Coarctation of the Aorta*

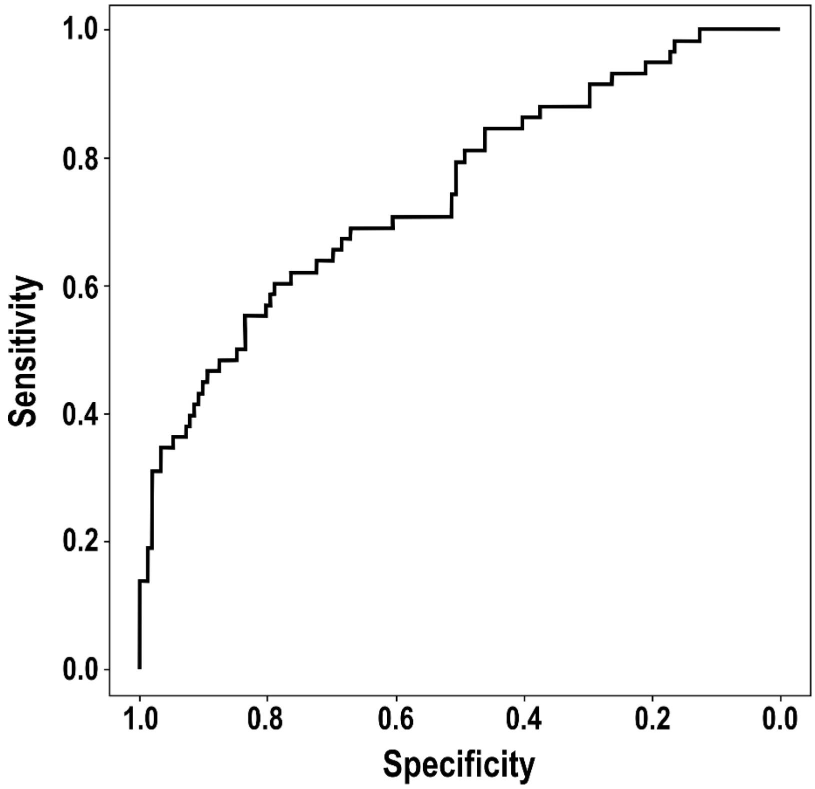


*Figure S2. ROC curve of the best model tested using retrospective data from treated CoA patients. Age at surgery and follow-up time since surgery were associated with hypertension, as well as four pairwise interactions: age and BPGpp at surgery, age at surgery and body surface area, follow-up time since surgery and body surface area, and follow-up time since surgery and hypertensive medication. Applying a cutoff of 22% with the model confusion matrix resulted in 72% sensitivity and 57% specificity and 0.74 area under the curve.*
